# Handling and accuracy of four rapid antigen tests for the diagnosis of SARS-CoV-2 compared to RT-qPCR

**DOI:** 10.1101/2020.12.05.20244673

**Authors:** Flaminia Olearo, Dominik Nörz, Fabian Heinrich, Jan Peter Sutter, Kevin Rödel, Alexander Schultze, Julian Schulze Zur Wiesch, Platon Braun, Lisa Oesterreich, Benno Kreuels, Dominic Wichmann, Martin Aepfelbacher, Susanne Pfefferle, Marc Lütgehetmann

## Abstract

**Background:** SARS-CoV-2 molecular diagnostics is facing material shortages and long turnaround times due to exponential increase of testing demand.

**Objective:** We evaluated the analytic performance and handling of four rapid Antigen Point of Care Tests (AgPOCTs) I-IV (Distributors: (I) Roche, (II) Abbott, (III) MEDsan and (IV) Siemens).

**Methods:** 100 RT-PCR negative and 84 RT-PCR positive oropharyngeal swabs were prospectively collected and used to determine performance and accuracy of these AgPOCTs. Handling was evaluated by 10 healthcare workers/users through a questionnaire.

**Results:** The median duration from symptom onset to sampling was 6 days (IQR 2-12 days). The overall relative sensitivity was 49.4%, 44.6%, 45.8% and 54.9 % for tests I, II, III and IV, respectively. In the high viral load subgroup (containing >10^6^ copies of SARS-CoV-2 /swab, n=26), AgPOCTs reached sensitivities of 92.3% or more (range 92.3%-100%). Specificity was 100% for tests I, II and IV and 97% for test III. Regarding handling, test I obtained the overall highest scores, while test II was considered to have the most convenient components. Of note, users considered all assays, with the exception of test I, to pose a significant risk for contamination by drips or spills.

**Discussion:** Besides some differences in sensitivity and handling, all four AgPOCTs showed acceptable performance in high viral load samples. However, due to the significantly lower sensitivity compared to RT-qPCR, a careful consideration of pro and cons of AgPOCT has to be taken into account before clinical implementation.

## Introduction

The SARS-CoV-2 pandemic continues to spread at accelerating pace [1], posing an unprecedented challenge for health care systems around the globe. Reverse transcription-quantitative polymerase chain reaction (RT-qPCR) analysis remains the gold standard for diagnosis of SARS-CoV-2 infection [2]; however, limited supply of material and specialized personnel, as well as frequent reporting delays bring about the need for alternative testing formats. Rapid Antigen Point of Care Tests (AgPOCTs) represent one such alternative and have been pushed into the market by countless distributors in recent months. The tally of CE-cleared AgPOCT products has mounted to just over 200 during this time [3], leaving potential users mostly in the dark about their performance in real-world situations as independent comparison studies were unable to keep pace with the flood of novel tests [4,5].

## Objective

The aim of this study was to evaluate and compare handling and analytical performance of four commercial rapid antigen point of care tests (AgPOCT) distributed by established diagnostics firms.

## Methods

### Clinical samples and ethics

Single center non-interventional study. Respiratory samples were oropharyngeal swabs collected using UTM based collection kits by Copan (Italy, Brescia) or Iclean (Shenzhen, China) and prospectively collected following routine diagnostics at the University Medical Center Hamburg-Eppendorf (during August through November 2020). This work was conducted in accordance with §12 of the Hamburg hospital law (§12 HmbKHG). The use of anonymized samples was approved by the ethics committee, Freie und Hansestadt Hamburg, PV5626.

### AgPOCT and RT-qPCR procedures

Four different CE-labeled AgPOCT kits were compared (Table 1). Swabs supplied with the AgPOCT kits were immersed in patient oropharyngeal samples for approximately 10 seconds before tests were carried out according to instructions of the manufacturer. Based on previous experience, we postulated that roughly 100µL of UTM sample are carried over by this procedure. To allow for a meaningful clinical correlation, ‘effective’ viral loads/swab were calculated by applying a factor of 0.1 to all PCR measurements from original samples. For limit of detection experiments, a serial dilution of cell-free virus solution containing SARS-CoV-2 strain HH-1 [6] in UTM was prepared and subjected to all four tests with five repeats per dilution step. As RT-qPCR reference method, the cobas6800 SARS-CoV-2 IVD assay was used in conjunction with quantitative external control material by Instand e.V. (Düsseldorf, Germany) to allow for absolute quantification, as previously described [7,8]. To evaluate the ease of handling and implementation of AgPOCTs into clinical routine we conducted a user survey employing a questionnaire (see supplementary Table S1) with 10 participants representing different clinical specialties and professions (3 ICU medical doctors, 2 ICU nurses, 3 microbiologists and 2 lab technicians). Every user received the complete set of materials for every test, including the instructions manual. Questionnaires were filled in anonymously and independently by each participant. A 5-point Likert scale (1 = “do not agree at all” to 5 = “absolutely agree”) was used to evaluate for different test dimensions. Statistical analysis was performed with STATA (version 15) and GraphPad Prism (version 86 9.0.0).

**Table 1.**
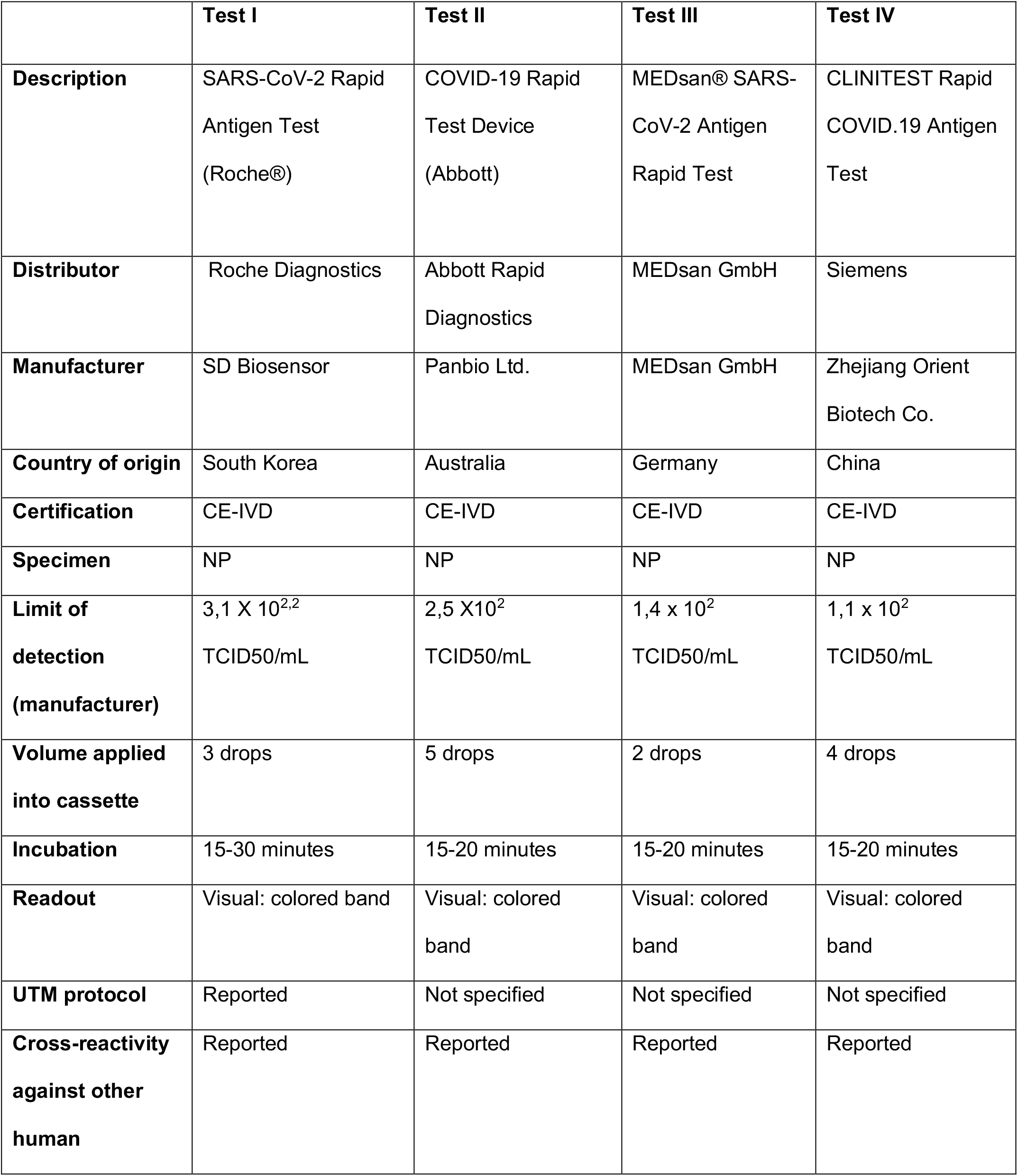
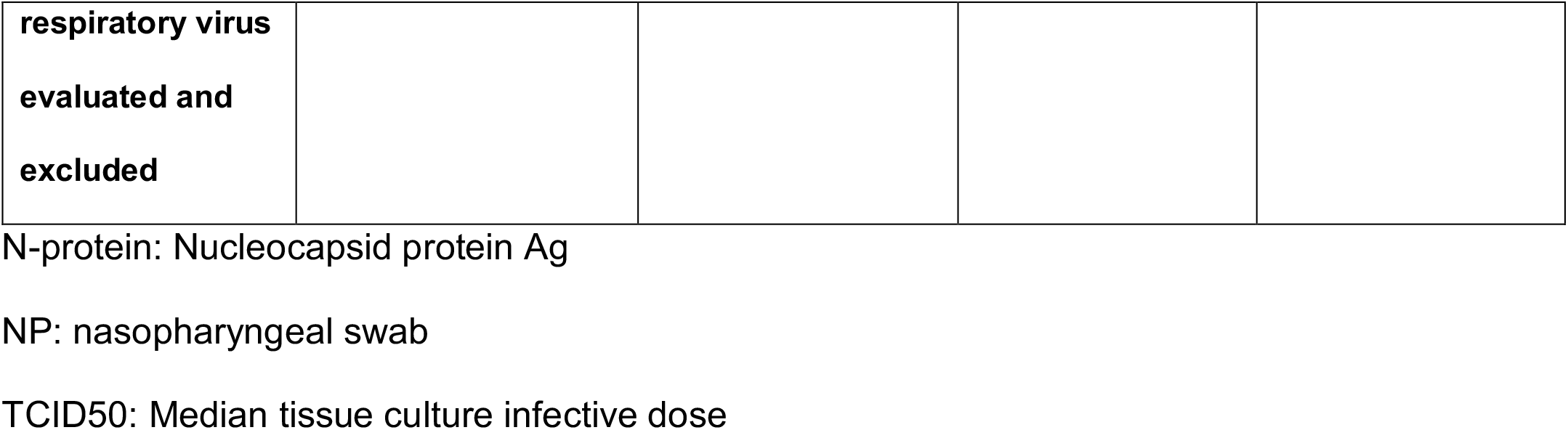
Main characteristics of four rapid SARS-CoV-2 rapid antigen tests (according to the manufacturer)

## Results

### Analytical and clinical performance

In total 100 RT-qPCR negative and 84 positive respiratory samples were included in this study. The median duration from symptom onset to sampling was 6 days (IQR 2-12 days). Consequently, 56.5% (26/ 46, unknown 38) of specimens were taken within one week of symptom onset. Median viral load/ swab for the AgPOCT was 3.8 x 10^5^ copies/swab (IQR 5.4 x 10^3^ – 3.8 x 10^6^ copies/swab). Clinical sensitivity was 49.4% (CI95: 38.9%-59.9%), 44.6% (CI95: 34.3% - 55.3%), 45.8% (CI95: 35.5% - 56.5%) and 54.9 % (CI95: 43.4% - 65.9%) for the tests I, II, III and IV, respectively (Table 2). LOD using cell culture derived SARS-CoV-2 virus was in line with these results showing similar analytical performance with best performance for test IV (see suppl. Figure S1). All assays showed a specificity of 100%, with the exception of test III (97% (CI95: 91.5% - 98.9%), see table 2). In a set of samples that all contained > 10^6^ copies/swab, clinical sensitivity rose to 100% (CI95: 87% −100%), 92.3% (CI95: 75.8% - 97.8%), 92.3% (CI95: 75.8% - 97.8%) and 100% (CI95: 85.7% −100%) for tests I, II, III and IV, respectively.

**Table 2.** Performance of four rapid antigen tests for SARS-CoV-2 compared to RT-qPCR.

| **Test** | **Sensitivity**<br>% (CI95) | **Specificity**<br>% (CI95) | **Accuracy**<br>% | **Kappa**<br>**coefficient** |
| --- | --- | --- | --- | --- |
| **I** (n=184) | 49.4 (38.9-59.9) | 100 (96.3 - 100) | 77.1 | 0.52 |
| **II** (n=184) | 44.6 (34.3-55.3) | 100 (96.3 - 100) | 74.8 | 0.46 |
| **III** (n=184) | 45.8 (35.5-56.5) | 97 (91.5 - 98.9) | 73.7 | 0.45 |
| **IV** (n=170) | 54.9 (43.4-65.9) | 100 (96.3 - 100) | 81.2 | 0.58 |

### Handling

The median of the results for each item is illustrated in figure 3. Test I was considered the overall easiest to use while test II had the easiest to use test components. Test III scored lowest overall. Of note, users considered all assays, with the exception of test I, to pose a significant risk for contamination by drips or spills (see Figure 2).

**Figure 1.**
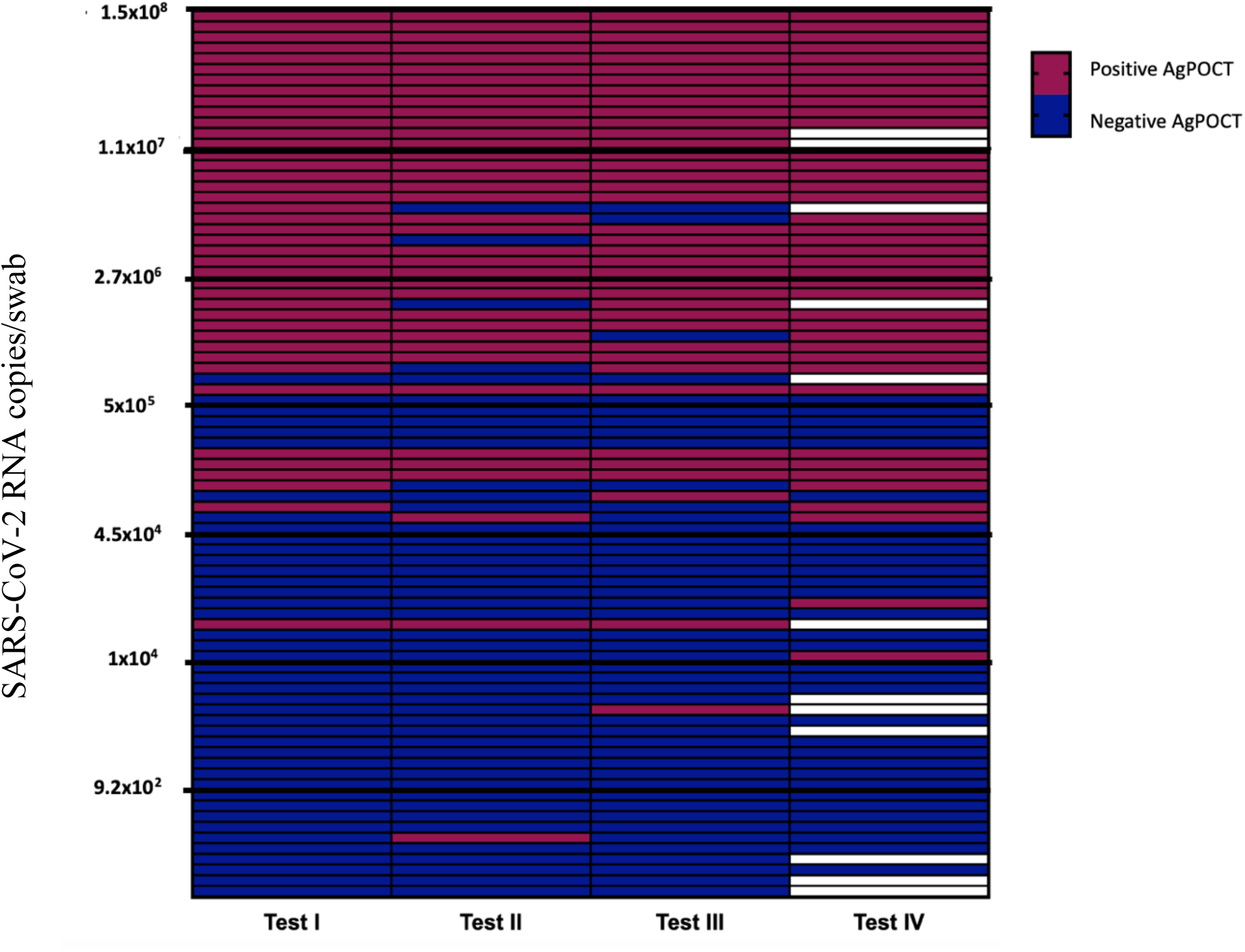
AgPOCT results in relation to SARS-CoV-2 RNA copies/swab. Purple fields represent positive and blue fields negative AgPOCT results. Blank fields correspond to cases where samples were not tested for the corresponding assay.

**Figure 2.**
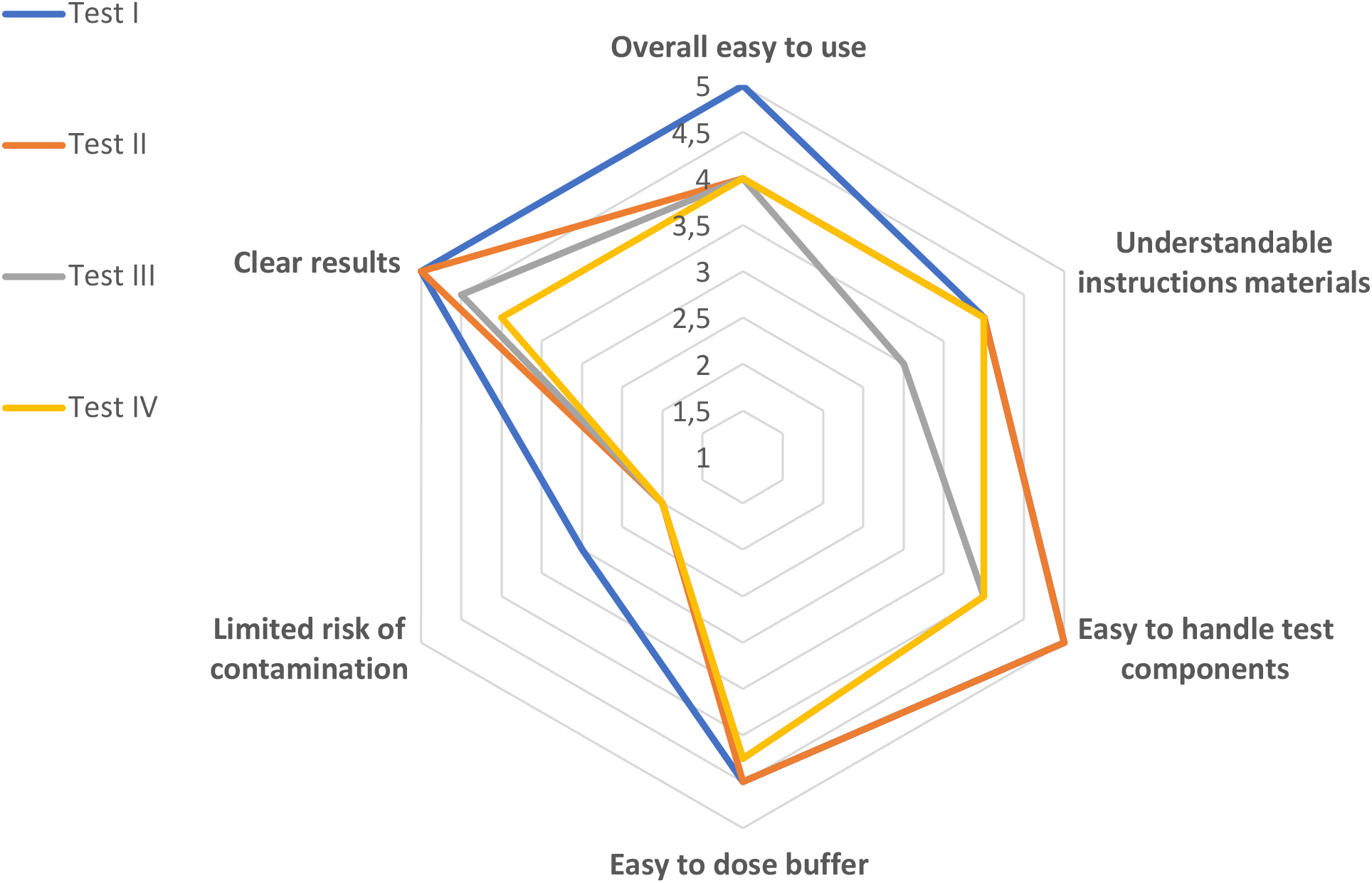
Evaluation of the handling of four AgPOCTs. Scores vary from 1 (“do not agree at all”) to 5 (“absolutely agree”).

## Discussion

Here we report a head-to-head comparison of performance and handling for four rapid AgPOCTs of several major distributors in Europe, using the latest version of CE-labeled kits. Relative clinical sensitivity of rapid SARS-CoV-2 antigen tests has repeatedly fallen short of optimistic manufacturer claims [4,9–11], in large parts due to differences in patient characteristics in respective cohorts and artificial conditions when performing AgPOCT. This also applies to this study, as e.g. antigen tests were performed from clinical UTM samples. Still, the lack of an amplification step inherently entails significantly higher analytic limits when compared to RT-qPCR, which is expected to be 100 – 1000 times more sensitive than an AgPOCT. However, in the context of increased frequency of testing, lower sensitivity tests can be just as effective as highly sensitive ones in identifying individual infections and outbreaks [12].

All AgPOCT evaluated in this study showed largely comparable clinical and analytical sensitivity, with the exception of Test IV (Siemens). The slight edge Test IV had in LoD experiments translated into a moderately increased positive agreement with RT-qPCR (of 54%) compared to the other tests (lower than 50%). It has to be noted that less clinical samples were tested with Test IV as it only became available when experiments were already underway, thus introducing a bias.

All tests were able to detect 10^6^ or more copies/swab with high reliability (95%), implying that patients with high viral loads can be identified with acceptable accuracy. This is in line with previous studies [4,5].

Besides analytical properties, handling and practical application was evaluated as part of this study. While all AgPOCTs were deemed “easy to use”, Test I was considered slightly better than the rest, while Test II was considered to have the most convenient components. We were surprised to notice that most users reported a high risk of contamination (by drips or spills). This aspect should not be underestimated in particular considering AgPOCT are to be used outside the health care sector and might also be performed by untrained personnel lacking experience in dealing with infectious materials. Especially in a context where test performance is largely identical, practical aspects should not be neglected when choosing which test to employ, as minor inconveniences in performing a single test can add up to massive delays when performing them in large numbers.

In conclusion, we present analytical and clinical evaluation of four rapid SARS-CoV-2 antigen tests, as well as a user survey for practical application. All tests demonstrated largely similar performance and are expected to detect high viral loads with adequate reliability. RT-qPCR remains the gold standard to definitively confirm or rule out infections due to its significantly higher sensitivity and specificity.

## Data Availability

Not applicable

## Abbreviations

RT-qPCR: Reverse transcription-polymerase chain reaction
AgPOCT: 

## Bibliography

[1] Covid-19: Is a second wave hitting Europe? | The BMJ, (n.d.). https://www.bmj.com/content/371/bmj.m4113 (accessed November 19, 2020).

[2] Laboratory testing for 2019 novel coronavirus (2019-nCoV) in suspected human cases, (n.d.). https://www.who.int/publications-detail-redirect/10665-331501 (accessed November 19, 2020).

[3] Liste der Antigentests, (n.d.). https://antigentest.bfarm.de/ords/antigen/r/antigentests-auf-sars-cov-2/liste-der-antigentests?session=21375807250974 (accessed November 24, 2020).

[4] T. Weitzel, P. Legarraga, M. Iruretagoyena, G. Pizarro, V. Vollrath, R. Araos, J.M. Munita, L. Porte, Head-to-head comparison of four antigen-based rapid detection tests for the diagnosis of SARS-CoV-2 in respiratory samples, BioRxiv. (2020) 2020.05.27.119255. https://doi.org/10.1101/2020.05.27.119255.

[5] Comparison of seven commercial SARS-CoV-2 rapid Point-of-Care Antigen tests | medRxiv, (n.d.). https://www.medrxiv.org/content/10.1101/2020.11.12.20230292v1.article-info (accessed November 24, 2020).

[6] S. Pfefferle, S. Reucher, D. Nörz, M. Lütgehetmann, Evaluation of a quantitative RT-PCR assay for the detection of the emerging coronavirus SARS-CoV-2 using a high throughput system, Eurosurveillance. 25 (2020). https://doi.org/10.2807/1560-7917.ES.2020.25.9.2000152.

[7] D. Nörz, N. Fischer, A. Schultze, S. Kluge, U. Mayer-Runge, M. Aepfelbacher, S. Pfefferle, M. Lütgehetmann, Clinical evaluation of a SARS-CoV-2 RT-PCR assay on a fully automated system for rapid on-demand testing in the hospital setting, J. Clin. Virol. Off. Publ. Pan Am. Soc. Clin. Virol. 128 (2020) 104390. https://doi.org/10.1016/j.jcv.2020.104390.

[8] D. Nörz, A. Frontzek, U. Eigner, L. Oestereich, D. Wichmann, S. Kluge, N. Fischer, M. Aepfelbacher, S. Pfefferle, M. Lütgehetmann, Pushing beyond specifications: Evaluation of linearity and clinical performance of the cobas 6800/8800 SARS-CoV-2 RT-PCR assay for reliable quantification in blood and other materials outside recommendations, J. Clin. Virol. 132 (2020) 104650. https://doi.org/10.1016/j.jcv.2020.104650.

[9] G.C. Mak, P.K. Cheng, S.S. Lau, K.K. Wong, C. Lau, E.T. Lam, R.C. Chan, D.N. Tsang, Evaluation of rapid antigen test for detection of SARS-CoV-2 virus, J. Clin. Virol. 129 (2020) 104500. https://doi.org/10.1016/j.jcv.2020.104500.

[10] A. Scohy, A. Anantharajah, M. Bodéus, B. Kabamba-Mukadi, A. Verroken, H. Rodriguez-Villalobos, Low performance of rapid antigen detection test as frontline testing for COVID-19 diagnosis, J. Clin. Virol. Off. Publ. Pan Am. Soc. Clin. Virol. 129 (2020) 104455. https://doi.org/10.1016/j.jcv.2020.104455.

[11] J. Dinnes, J.J. Deeks, A. Adriano, S. Berhane, C. Davenport, S. Dittrich, D. Emperador, Y. Takwoingi, J. Cunningham, S. Beese, J. Dretzke, L. Ferrante di Ruffano, I.M. Harris, M.J. Price, S. Taylor-Phillips, L. Hooft, M.M. Leeflang, R. Spijker, A. Van den Bruel, Cochrane COVID-19 Diagnostic Test Accuracy Group, Rapid, point-of-care antigen and molecular-based tests for diagnosis of SARS-CoV-2 infection, Cochrane Database Syst. Rev. 8 (2020) CD013705. https://doi.org/10.1002/14651858.CD013705.

[12] M.J. Mina, R. Parker, D.B. Larremore, Rethinking Covid-19 Test Sensitivity — A Strategy for Containment, N. Engl. J. Med. (2020). https://doi.org/10.1056/NEJMp2025631.

