## Supplemental Figure S1 and Table S1 for "Handling and accuracy of four rapid antigen tests for the diagnosis of SARS-CoV-2 compared to RT-qPCR": Supplementary material.docx

**Figure S1.** SARS-CoV-2 isolate HH-1 (6) was quantified by plaque assay as described (8). Serial dilutions of virus were prepared in UTM and swabs provided by the AgPOCT manufacturer were dipped into the virus containing solution. Thereafter, assays (test I in green, test II in blue, test III in grey and test IV in red) were performed as recommend by the manufacturer.

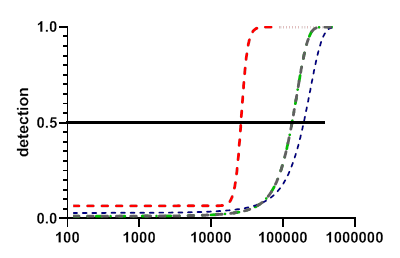

**PFU/ml**

**Table S1.** Questionnaire on AgPOCT handling.

**ITEM 1: Overall easy to use**

- Overall, the test execution is simple.

**ITEM 2: Understandable instruction materials**

- The manual instruction is easy to understand
- The manual instruction is exhaustive
- The short manual is useful

**ITEM 3: Easy to handle test components**

- The test material contained is clearly marked
- The test material is easy to open
- The test material is easy to use

**ITEM 4: Easy to dose buffer**

- The Buffer is easy to apply
- The dosage of the buffer can be carried out reliably and precisely

**ITEM 5: Limited risk of contamination**

- The implementation of the assay doesn´t bring to risk of contamination (e.g. by dripping / spilling)

**ITEM 6: Clear results**

- The test result can be read clearly
